# The Intrathecal Morphine for Percutaneous Endoscopic Lumbar Discectomy (IMPELD) Study: Rationale and Protocol for a Double-blinded Randomized Placebo-controlled Trial

**DOI:** 10.1101/2021.12.08.21267387

**Authors:** Yue Lei, Zhang Feng, Mu Guanzhang, Shang Meixia, Sun Haolin, Lin Zengmao

## Abstract

**Background:** Percutaneous endoscopic lumbar discectomy (PELD), a minimally invasive spinal technique for lumbar disc herniation (LDH), has gained popularity globally and yielded satisfying results. However, PELD is often performed on awaking patients to avoid nerve injury, thus the intraoperative analgesia of PELD is sometimes insufficient. The effect of intrathecal morphine (ITM) has been well proved in various surgical specialties, and this study aims to investigate the effectiveness and safety of ITM on PELD.

**Methods:** The intrathecal morphine for percutaneous endoscopic lumbar discectomy (IMPELD) trial is a double-blind, randomized, placebo-controlled trial. The 90 eligible LDH patients undergoing PELD will be randomly assigned to receive either ITM or placebo during spinal anesthesia, at a 1:1 ratio, with a one-month follow-up period. Average intraoperative pain intensity will be the primary outcome. Secondary outcome measures include intraoperative pain intensity assessed at each 30 min intraoperatively, postoperative pain intensity, perioperative analgesia requirements, functional evaluation, radiographic characteristics, overall satisfaction, other characteristics and adverse events.

**Discussion:** Currently, there is a lack of scientific evidence to provide a reliable method to reduce intraoperative pain of PELD. The IMPELD trial was designed to provide evidence regarding whether 100 ug of ITM is an effective and safe coanalgesic approach for PELD procedure.

**Trial registration:** The trial was registered with the Chinese Clinical Trial Registry (identifier ChiCTR2000039842). Registered on November 11^th^, 2020.

## 1. Background

Lumbar disc herniation (LDH) is a common cause of sciatica affecting one’s performance at work and quality of life.[1] Since the introduction of the percutaneous endoscopic lumbar discectomy (PELD) in the 1990s, this minimally invasive technique has becoming an idealizing alternative for LDH for its less surgical trauma, faster postoperative recovery and fewer complications.[2] Given the technical improvements of endoscopic spine surgery, including optics design, surgical instruments, and specific surgical approach, its clinical application is becoming increasingly widespread and standardized.[3, 4] PELD is often performed under conscious state, i.e., local, intrathecal or epidural block, rather than under general anesthesia, to prevent potential nerve damage during the operation [5]. However, for the awaking patients, the pain during the PELD operation is sometimes difficult to endure and might lead to severe sequelae, such as post-traumatic stress disorder [6, 7]. An effective and safe supplementary perioperative analgesia for PELD procedure is therefore in need.

A potential adjuvant approach would be intrathecal morphine (ITM), which was firstly introduced for pain relief in clinical in 1979, and since then its efficacy has been well established in various surgical scenarios.[8-11] As the subarachnoid space is readily accessible during spinal anesthesia, it seems reasonable that ITM could be used for perioperative analgesia in patients undergoing PELD.

We here report the clinical and statistical design of the intrathecal morphine for percutaneous endoscopic lumbar discectomy (IMPELD) trial, a randomized, double-blind, placebo-controlled trial in LDH patients. The study aims to investigate the effectiveness and safety of ITM on an endoscopic spinal procedure. It is hypothesized that ITM as an adjuvant analgesic would significantly reduce the intraoperative pain intensity during PELD.

## 2. Methods/design

### 2.1 Participants

This trial will be conducted at Peking University First Hospital, involving LDH patients eligible for single-space PELD by a single well-trained surgeon (SHL). Inclusion criteria: (1) At least 18 years of age; (2) radiculopathy due to disc herniation, defined as having 2 or more of the symptoms or physical tests (i.e., the straight leg raising test, crossed straight leg raising test, paresis or muscle weakness, sensory deficits, or impaired reflexes), confirmed by magnetic resonance imaging (MRI) [12]; (3) who choose to undergo a single segment PELD under intrathecal anesthesia; (4) ASA score ≤ 3. The exclusion criteria are as follows: (1) morphine use history within 3 days; (2) contraindications of morphine use or lumbar puncture; (3) respiratory disorders, e.g., obstructive sleep apnoea syndrome (OSAS), chronic obstructive pulmonary disease (COPD), asthma, or chronic cough; (4) obese patients (BMI >= 30); (4) mental disorders, cognitive impairment, unable to cooperate with pain assessment, illiteracy, deaf/mute, or drug addicts; (5) pregnant and lactating women.

All participants will sign an informed consent. The study was approved by the Institution Review Board of Peking University First Hospital (2020-289) and prospectively registered at Chinese Clinical Trial Registry (ChiCTR2000039842).

### 2.2 Randomization and blinding

The eligible patients will be randomly assigned into either the ITM or the control group with an equal allocation ratio (1:1) with a block size of 4 by an independent biostatistician (SMX) using SAS statistical package version 9.4 (SAS Institute, Cary, NC, USA). Each patient’s allocation will be concealed in a sequentially numbered, sealed, opaque envelope. The envelope will be kept and opened by a senior anesthetist (LZM), the only researcher knowing the allocation, when a patient arrives at the anesthetic induction room. The patients, surgeons, investigators, nurses and the statistician in the whole process will be all blinded to group allocations.

Besides, patients who wish to participate in the trial but do not consent to randomization or patients who do not meet eligibility criteria will be included in a nonrandomized observational cohort, considered as equivalent to the control group, and will be followed up.

### 2.3 Interventions

Before lumbar puncture, the back skin is disinfected, draped, and infiltrated with 2 ml of 1% lidocaine for local anesthesia in both groups. The subarachnoid space puncture is performed with a 25-gauge pencil-point spinal needle (Tuoren, Xinxiang, China), with the following confirmation of the return of clear, free-flowing cerebrospinal fluid. All patients will receive a routine 6 ml of hypobaric ropivacaine (0.125%) injection for sensorial block. Next, 100 μg of morphine diluted in 2 ml of 0.9% saline solution will be administered intrathecally over 10 seconds in the ITM group, while 2 ml of 0.9% saline solution administered at the same velocity in the control group. Then patient lie in prone position and the PELD procedure is performed. At 1h after returning to the ward, 100 mg of flurbiprofen axetil (diluted with 100 ml of 0.9% saline solution) will be routinely administered in all subjects for post-operative analgesia. Patients will be routinely discharged at 24 h after PELD, if there are no special circumstances. The flow of the study is illustrated in Figure 1.

**Figure 1.**
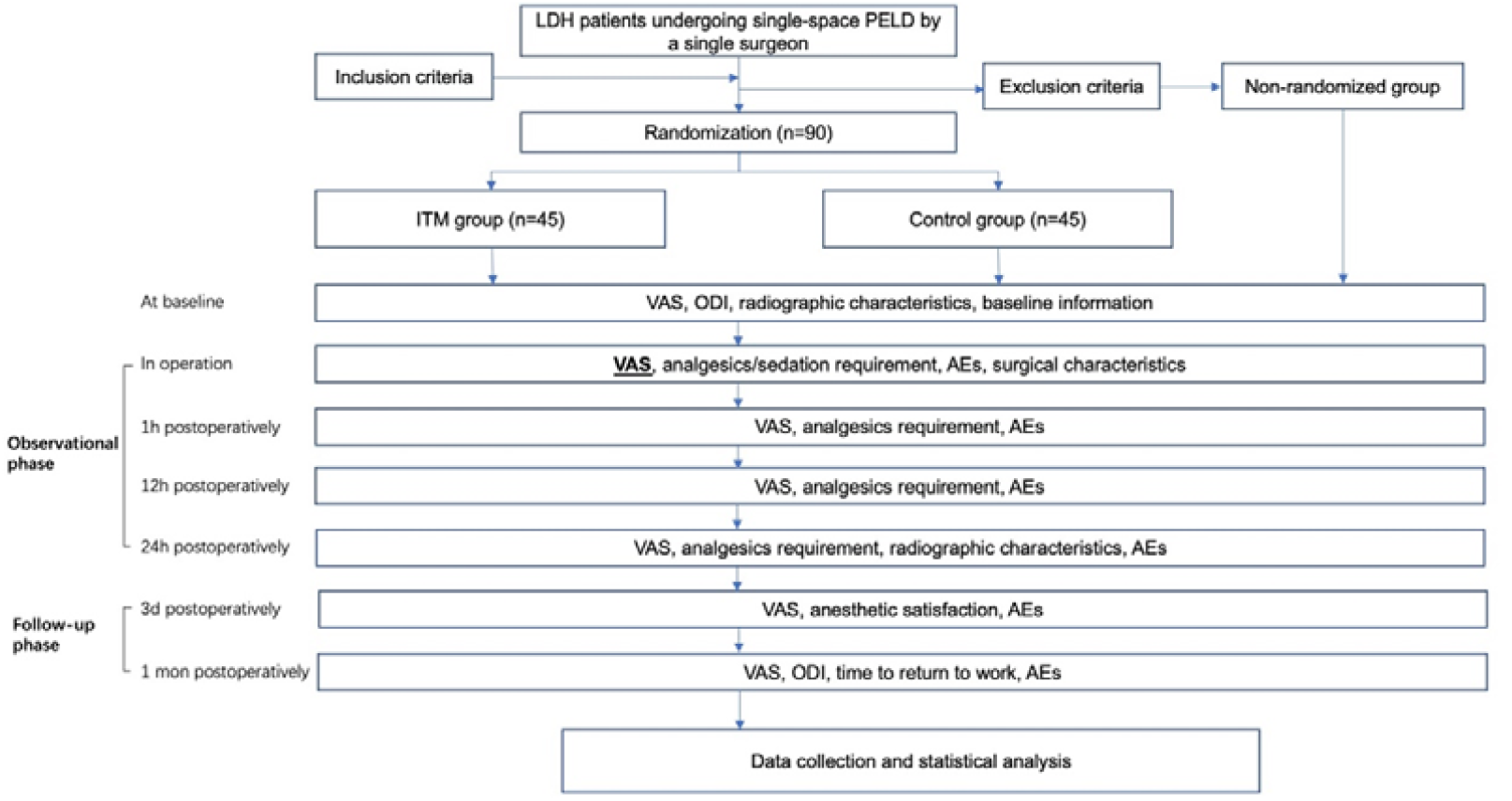
Schematic flow of patient flow. *AE = adverse event, ODI = Owestry Disability Index, VAS = visual analogue scale.*

### 2.4 Outcomes

#### 2.4.1 Primary outcome

The primary outcome is the average intra-operative self-reported pain intensity evaluated on a 10-point visual analogical scale (VAS) scale right after the operation ended. The VAS scores will be collected by a blinded investigator (YL), for details see supplementary information.

#### 2.4.2 Secondary outcomes

##### (1) Intra-operative self-reported pain intensity

the intra-operative pain intensity will be evaluated the following time-points: at the time canula inserted and at every 30 min afterwards until the end of the operation (e.g., 30 min after canula inserted, 60 min after canula inserted and so on). It should be noted that the intra-operative pain intensity will be only evaluated as surgical site pain, as surgical instruments may cause transient radicular symptoms intraoperatively.

##### (2) Post-operative self-reported pain intensity

the post-operative pain intensity will be evaluated both during hospital stay (1 h, 12 h, and 24 h post-operatively) and after hospital discharge (3 day postoperatively and 1 month postoperatively). The pain intensity of pain will be evaluated at the both back (or surgical site) and lower extremity, both at static and during movement.

##### (3) Analgesic requirements

The number of patients requiring analgesia and/or sedation during surgery. Besides, the number of patients in need of additional analgesics after returning to the ward will also be recorded.

##### (4) The functional evaluation

Owestry Disability Index (ODI) will be used for evaluating the disability score at baseline and at 1 month follow-up. Time to return to work will also be documented.

##### (5) Radiographic parameters

Sagittal parameters and coronal parameters of the spine on X-ray in upright position will be collected and measured before surgery and 24 h after surgery. Baseline MRI characteristics include protrusion size & zone classification[13], disc degeneration grade[14], facet joint degeneration grade[15], and presence of special changes (Modic change, Schmorl’s node, high intensity zone) [16-18]. All radiographic parameters will be assessed by two independent raters (YL and MGZ). Numerical results of two raters will be averaged. For grade variables, a consensus-based decision will be adopted if there is an interrater disagreement.

##### (6) Overall satisfaction

the overall anesthetic satisfaction will be self-reported by the 5-point Likert scale at 72 h postoperatively.

##### (7) Other characteristics

Operation duration, approach of PELD (transforaminal or interlaminar), and whether foraminoplasty is performed or not will be obtained from the medical record.

##### (8) Adverse events

Any AEs occurring during the whole follow-up period will be recorded. We here predefine the ITM-related adverse events as respiratory depression, nausea or vomiting, pruritus, hypotension or urinary retention[19, 20]. Respiratory depression is predefined as oxygen saturation less than 90% in a 30 second window regardless of symptoms, after excluding instrument error[21]. Hypotension is predefined as the systolic blood pressure less than 90 mm Hg for a least 5 mins[22]. Other adverse events, such as anesthetic complications, surgical complication, re-admission, reoperation, etc., will also be documented. What’s more, the severity of AEs is graded from 0 to 5, and AEs of grade 3, grade 4 and grade 5 will be defined as severe adverse events (SAEs) in the current study, see Table 1[23]. The full list of assessment at each time-point is detailed in Table 2.

**Table 1.** Common Terminology Criteria for Adverse Events[23]. *ADL = Activities of Daily Living*.

| Grade | Definition |
| --- | --- |
| 0 | None |
| 1 | **Mild**; asymptomatic or mild symptoms; clinical or diagnostic observations only; intervention not indicated. |
| 2 | **Moderate**; minimal, local or noninvasive intervention indicated; limiting age-appropriate instrumental ADL |
| 3 | **Severe** or medically significant but not immediately life-threatening; hospitalization or prolongation of hospitalization indicated; disabling; limiting self-care ADL |
| 4 | **Life-threatening** consequences; urgent intervention indicated. |
| 5 | **Death** related to AE |

**Table 2.** List of assessment at each time-point.

|  | Screening | Observation |  |  |  | Follow-up |  |
| --- | --- | --- | --- | --- | --- | --- | --- |
| Timepoint | Baseline | Intraoperative | 1 h postoperatively | 12 h postoperatively | 24 h postoperatively | 72 h postoperatively | 1 mon postoperatively |
| Baseline characteristics | ♦ |  |  |  |  |  |  |
| Pain intensity | ♦ | ♦ | ♦ | ♦ | ♦ | ♦ | ♦ |
| Radiography | ♦ |  |  |  | ♦ |  |  |
| Analgesia (or sedation) requirements |  | ♦ | ♦ | ♦ | ♦ |  |  |
| Functional evaluation | ♦ |  |  |  |  |  | ♦ |
| Adverse events |  | ♦ | ♦ | ♦ | ♦ | ♦ | ♦ |
| Overall satisfaction |  |  |  |  |  | ♦ |  |
| Other characteristics |  | ♦ |  |  |  |  |  |

### 2.5 Reporting and treating adverse events

The investigator may unblind the treatment allocation of a participant, in the presence of severe adverse events (AEs) for performing immediate remedy, by asking the senior anesthetist. Unanticipated problems related to participation in the research will be reported to the IRB.

In prevention of potential adverse events during operation, routine monitoring will be commenced (pulse oximetry, noninvasive blood pressure monitoring and electrocardiography), and intravenous access will be established. For preventing post-operative AEs, monitoring will be performed within the 12 h post-operatively for all patients. Naloxone will be routinely prepared for each subject in case of opioid-related side-effects needing to be rescued.

### 2.6 Sample size calculation

The pilot study of 10 patients showed that the average intra-operative VAS in ITM group was 0.60 ± 0.55 and the VAS in the Control group was 1.00 ± 0.71. Considering a drop-out rate of 10% (including failure to perform spinal anesthesia), a sample size of 45 patients per group would be sufficient in this study with a statistical power of 0.8 and a false-positive error rate of P-value ≤ 0.05 on a 2-tailed Student t test.

### 2.7 Data management and monitoring

All experimental data will be firstly entered into a paper-based case report form (CRF) by a blinded investigator (YL). Participant identification and privacy information will be de-identified to prevent participants’ personal identity from being revealed. All outcomes on paper CRF will be independently imputed using EpiData 3.1 (EpiData Association, Odense, Denmark) by another blinded investigator (MGZ), after which the informed consent and completed questionaries will be kept in locked cabinets/rooms only accessible by the biostatistician (SMX), the only researcher with access to the final trial dataset after data storage. Drop-outs and withdrawals from the trial will be recorded. Lastly, treatment allocation of all participants will be unblinded for analysis purposes at the end of the trial.

### 2.8 Statistical methods

Data will be analyzed using the statistical software package SPSS version 28.0 (Chicago, IL, USA). All analyses will be on an intention-to-treat (ITT) basis. Continuous variables will be described as the mean ± standard deviations (SDs). An independent-samples t-test will be used for continuous variables with a normal distribution, and the Mann–Whitney U will be used for data with a non-normal distribution. Categorical variables will be described as the number (%) and analyzed with the Fisher’s exact test or the chi-squared test. A two-sided P value of less than 0.05 was considered to be statistically significant.

For the primary outcome, post hoc subgroup analyses on the primary end points will be performed based on the following cut points: mean age, mean onset of symptoms, and gender. Besides, sensitivity analyses will be conducted by adding data from the non-randomized cohort, and excluding the data from patients requiring intraoperative analgesics and/or sedation.

The efficacy of blinding will be evaluated by the proportion of participants who believed they are treated with ITM (or active treatment), as will be reported at the last follow-up. If no significant difference is noticed, the blinding procedure will be considered successful. Multiple imputation approach will be applied for dealing with missing data.

## 3. Discussion

The key guarantee for the successful completion of PELD operation is to avoid nerve injury as the surgical instruments are performed around the dural sac/nerve root cuff during the procedure, and therefore PELD is typically performed local anesthesia or intraspinal anesthesia to retain consciousness. Previous evidence has shown that the intraspinal anesthesia showed superior analgesic efficacy to local anesthesia[6, 24]. However, intolerable pain could still occur in some patients with intraspinal anesthesia, while additional intravenous medication and subsequent complications related to sedatives and analgesics may be encountered. Therefore, a more effective and safe analgesia for PELD under intraspinal anesthesia is in urgent need.

Published literature provides only handful solutions to address this situation. Kim showed that intraoperative sedation by dexmedetomidine provides ideal sedative and hypnotic surgical condition on PELD, but high dexmedetomidine dosage resulted in delayed emergence [25]. The same scholar also reported that lidocaine patch before PELD resulted in better clinical efficacy compared with placebo, with only subtle adverse effects [5]. Another observation study by Fan et, al. demonstrated that intramuscular injection of morphine prior to PELD turns out to be overall effective, except for the increased risk of nausea and vomiting[26]. Our study is the first clinical trial to investigate the effect of ITM, a time-tested analgesia approach, as an adjuvant pain-relieving therapy on PELD.

ITM has been used for perioperative pain management since 1970s[8]. Effective analgesia of ITM can be obtained with doses ranging from 0.1 to 2.5 mg; however, recent studies have coincidentally preferred to investigate into loser dose of ITM for preventing potential opioids-related side-effects[19]. Although a recent RCT by Wang et, al showed that 200 ug of ITM on open lumbar surgery with general anesthesia results in significantly improved early postoperative pain relief and reduced postoperative analgesics consumption without increased risk of AEs, comparing with placebo[27]. However, the patients in our study will be under sober state without assisted ventilation, hence the risk of respiratory complications should be further minimized. Therefore, in order to ensure safety, the medication dose in the current study was set as 100 μg to maximally reduce the risks of opioids-related complications, especially respiratory depression. The preliminary trials of 5 patients undergoing ITM did not show notable AEs, except for a case of transient pruritus postoperatively. The rescue remedy for ITM is naloxone, the opioid antagonists to decrease opioid side effects. Therefore, we prepared naloxone routinely for each participant in cases of severe opioids-related complications.

Of a final note, if the hypothesis in the current study is proven, ITM could be recommended as an alternative analgesic approach for intraoperative analgesia in PELD.

### Trial status

The version number of this protocol is version 1.0, date: 2020-9-6. Recruitment started on 11 November 2020, and we expect to complete all research processes on May 2022.

## Data Availability

All data produced in the present study will be available at the end of the trial and will be available upon reasonable request to the authors.

## Supplementary information

Illustration figure of how the primary outcome will be assessed, via a third-party platform. http://doi.org/10.6084/m9.figshare.17150603.

## Authors’ contributions

Yue Lei, Lin Zengmao and Sun Haolin conceived of the study and designed the study protocol. Yue Lei and Zhang Feng drafted the manuscript and they contributed equally to the work. Yue Lei, Zhang Feng and Mu Guanzhang are in charge of coordination and direct implementation. Shang Meixia performed the randomization and allocation, and helped to develop the study outcomes and analyses. All authors contributed to drafting the manuscript and have read and approved the final manuscript.

## Acknowledgments

This study was funded by Interdisciplinary Clinical Research Project of Peking University First Hospital (2021CR31).

